# COVID-19 national lockdown in Morocco: impacts on air quality and public health

**DOI:** 10.1101/2020.07.05.20146589

**Authors:** Kenza Khomsi, Houda Najmi, Hassan Amghar, Youssef Chelhaoui, Zineb Souhaili

## Abstract

On the 20^th^ April 2020, the end date of the first strict lockdown period in Morocco, 2 403 410 cases of the corona Virus were confirmed globally. The number of Morocco confirmed cases attended 2990, while 12 746 were suspected and 143 deaths were recorded. Due to the pandemic of coronavirus disease 2019 worldwide and in Morocco, almost all avoidable activities in the country were prohibited since the kingdom announced activities reduction on March 16, 2020 and then general lockdown with reduced industrial activities on March 20, 2020.

This study aims at comparing the air quality status in Casablanca and Marrakech, two large cities from Morocco, before the pandemic and during the lockdown situation to show whether COVID-19 compelled-anthropogenic activities lockdown may have saved lives by restraining ambient air pollution than by preventing infection.

We found that, during the quarantine, NO_2_ dropped by -12 μg/m^3^ in Casablanca and -7 μg/m^3^ in Marrakech. PM_2·5_ dropped by -18 μg/m^3^ in Casablanca and -14 μg/m^3^ in Marrakech. CO dropped by -0.04 mg/m^3^ in Casablanca and -0.12 mg/m^3^ in Marrakech. This air pollution reduction had created human health benefits and had reduced mortality and saved lives mainly from cardiovascular diseases.

## 1. Introduction

Coronavirus disease 2019 (COVID-19) is an infectious disease initially identified in the city of Wuhan in China on the 31^st^ December 2019 (Khan and Naushad, 2020). On the 20^th^ April 2020, COVID-19 has led to more than 165 000 deaths worldwide with a global mortality rate of 6.8% (WHO, 2020a). Due to the contagious nature of COVID-19, most countries declared the lockdown, hence businesses and industrial activities decreased, global air travel reduced and cars and trucks stayed off the roads (Isaifan, 2020; Martins, 2020; Wang et al., 2020).

Morocco reported the first COVID-19 case on the 2^nd^ March 2020, 39 cases, and 1 death followed. Then the government announced the obligation of physical distancing and border closures on the 16^th^ March 2020 and gathering places closed. The lockdown followed on the 20^th^ March 2020. Since then, the unofficial transit going in and out of Moroccan cities was shut down, most transportation was prohibited and almost all avoidable outdoor human activities stopped all around the country. On the 20^th^ April 2020, the end date of the first strict lockdown period, Morocco announced 2990 confirmed cases and 143 deaths, thus a mortality rate of 4.8% (Ministry of Health, 2020).

Beyond the slowdown in the spread of the COVID-19 virus, the restrictive measures implemented by the authorities around the world have a second, less expected effect: that of reducing ambient air pollution which mortality rate has contributed to 7.6% of all deaths in 2016 worldwide (Isaifan, 2020). In China, Nitrogen Dioxide (NO_2_) and carbon emissions dropped by 30 and 25%, respectively (Isaifan, 2020). In India, the concentrations of Particulate Matter with a diameter of less than 10μm and 2.5μm (PM_10_ and PM_2.5_) reduced by 50% and NO_2_ has also shown considerable decline (Mahato et al., 2020). In Spain, the concentration levels of NO_2_ have declined by 64% in major cities (Universitat Politècnica de València, 2020). In Northern Italy, a drastic reduction in NO_2_ emissions was observed too (Martins, 2020). Emissions have also dropped in the USA (Newburger, 2020). In the city of Salé in Morocco, PM_10_ and NO_2_ concentrations dropped respectively by 75% and 96% during the period between the 11^th^ March and the 2^nd^ April 2020 (Otmani et al., 2020).

According to the World Health Organization (WHO), 4.2 million deaths have been caused by ambient air pollution worldwide in 2016. The latter is estimated to cause about 29% of lung cancer deaths, 24% of stroke deaths, 25% of heart disease deaths, and 43% of other lung diseases. Moreover, air pollution has attributed to 26% of respiratory infection deaths, 25% of chronic obstructive pulmonary disease (COPD) deaths, and about 17% of ischemic heart disease and stroke. It is noteworthy that chronic respiratory and cardiovascular diseases may be linked to COVID-19 as the death rate caused by the virus is higher among those with these diseases (Isaifan, 2020; WHO, 2020b). As for Morocco, each year more than 13,000 deaths is due to air pollution. This represents about 7% of all deaths thus the 8^th^ largest mortality risk factor. According to the global report “Toxic Air: The Price of Fossil Fuels” of the international NGO Greenpeace MENA, annually deaths due to air quality degradation in Morocco is estimated at 5,100 in 2018 (Farrow, A., Miller, K.A. & Myllyvirta, 2020).

Obviously, COVID-19 is a treat for public health, nevertheless, the accompanying lockdown measures, instigating improved air quality, may have positive effects on human life and well-being. For instance, Chen et. al estimate that improved air quality in China, during the quarantine period, avoided a total of 8911 NO_2_-related deaths and 3214 PM_2.5_-related deaths (Chen et al., 2020).

The present study has started in April 2020 to highlight the impacts of COVID-19 lockdown on air pollution levels in Morocco and the potentially avoided cause-specific mortality during population quarantine episode. We have adopted the same approach as (Chen et al., 2020). To the best of our knowledge, it reports on the first case study that links the COVID-19 to positive consequences on health in Morocco and tries to verify whether this infection has saved people than it has killed so far.

The remainder of this paper starts with presenting the materials and methods including the study area, the used data and, the adopted approach. It then presents the found results and the discussion section.

## 2. Materials and Methods

### 2.1. The study area

Morocco is an African country located in the extreme northwest of the continent (Fig. 1). It is located in the southern part of the Mediterranean basin and is considered among the most vulnerable countries to climate variability and transboundary air pollution (Khomsi et al., 2020). Casablanca and Marrakech are the studied cities. They are, respectively, the first and the fourth populous cities in Morocco with more than 3,000,000 and 900,000 residents, respectively, as reported by the World Population Review webpage (World Population Review, 2020). Casablanca is located in the central-western part of the country on the Atlantic Ocean coast (Fig. 1). In economic and demographic terms, Casablanca is one of the most important cities in Africa and is the largest city in the Maghreb. Marrakech is a large city in the Kingdom of Morocco. It is the chief town of the inland mid-southwestern region of Marrakech-Safi and located to the north of the piedmont of the snow-capped Atlas Mountains (Fig. 1). With the COVID-19 crisis, the cities of Casablanca and Marrakech account together for more than 50% of the people infected. On the 20^th^ April 2020, it has been reported that the region of Casablanca-Settat has reached exactly 840 cases leading the most regions affected by COVID-19, it is followed by the region of Marrakech-Safi which reached 739 cases of Coronavirus. Moreover, Casablanca and Marrakech are often chosen for air pollution researches as they are the urban areas where serious pollution concerns may be met, especially with the important population rate increase between 2004 and 2014, this rate reaches 11% in Casablanca and 12% in Marrakech.

**Figure 1:**
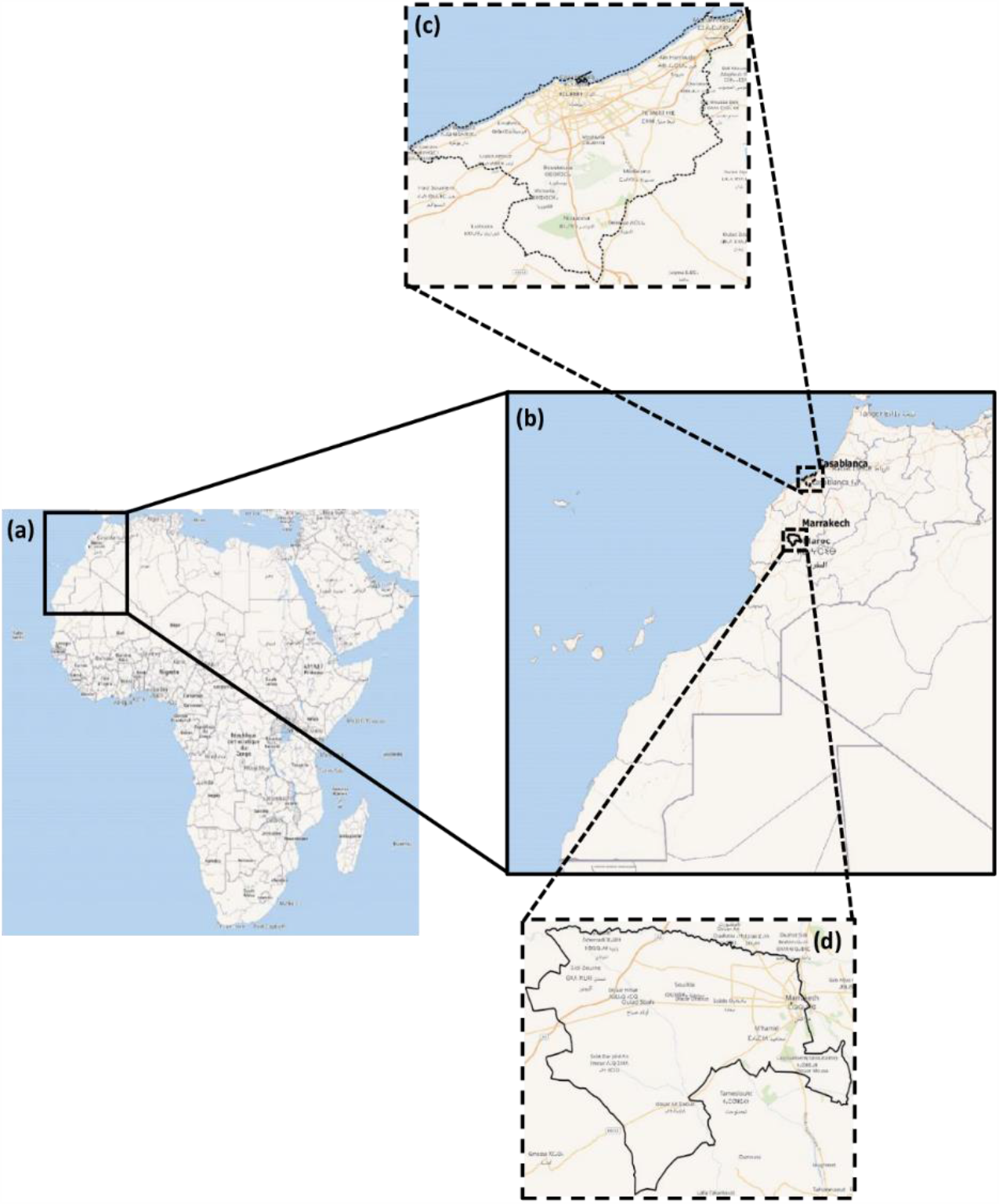
Location of the study area. (a) Africa, (b) Morocco, (c) Casablanca, (d) Marrakech

### 2.2. Air quality, health, and population data

The number of daily confirmed COVID-19 cases and death in Morocco were collected from the official website set by the ministry of health for that purpose (Ministry of Health, 2020). Air quality data (daily concentrations of NO_2_, PM_10_, and CO) recorded in air quality stations, were gathered from the National Weather Service in charge of air quality management in Morocco. PM_10_ concentrations were then converted to PM_2.5_ data for each station, using a conversion factor of 0.4 (Ait Bouh et al., 2013). We focused on NO_2_, PM_2·5_, and CO because they are traffic-related air pollutants whose emissions must obviously reduce as a result of the national traffic ban and home quarantine in Morocco during the lockdown.

Up to date city-level demographics are obtained from the World Population Review webpage (World Population Review, 2020). Country-level air quality related mortality including that incidental to chronic disease is collected from the Institute for Health Metrics and Evaluation (IHME) (IHME, 2017).

### 2.3. Methods

To assess air pollution reduction due to the lockdown, we used a difference-in-difference approach that we validated through the non-parametric approach proposed by Theil and Sen for univariate time series (Sen, 1968; Theil, 1950). Specifically, we defined the before quarantine period as between the 16^th^ February and the 19^th^ March 2020 and the during quarantine as between the 20^th^ March and 20^th^ April 2020. We assessed changes in air quality during vs. before the quarantine period in 2020 and compared these with corresponding changes in the same lunar calendar periods in 2016-2019. We assessed the statistical significance of the obtained trends using the modified Mann-Kendall test proposed by Hamed and Ramachandra Rao for autocorrelated time series (Hamed and Ramachandra Rao, 1998). The test is performed at a significance level of 5%.

In order to quantify the impact of the found change in air quality on health, we calculated the avoided cause-specific mortality attributable to the decreases in NO_2_ and PM_2.5_ over the study area based on the concentration-response functions from previous studies by (Chen et al., 2018, 2017) and the cause-specific mortality data from the IHME in 2017 (IHME, 2017). In addition to total non-accidental mortality, the cause-specific mortality for cardiovascular disease, hypertensive disease, stroke, chronic respiratory diseases, and chronic obstructive pulmonary disease (COPD) was also calculated. The attributable fraction (AF) method was used to estimate the daily avoided cause-specific mortality from air pollution reduction as done by (Chen et al., 2020). AF is defined as follows: *AF* = 1 − *e*^−*β*Δ*c*^ β is the cause-specific coefficient of the CRF, it is expressed as the percentage change in daily mortality associated with a 10 μg/m^3^ increase in daily NO_2_ or PM_2.5_. β values are available in the study by (Chen et al., 2020). Δc is the air quality changes due to the quarantine. AF is then multiplied by the daily cause-specific number of deaths and the total number of days during the quarantine period (32 days) to estimate the cause-specific avoided deaths.

## 3. Results and discussion

### 3.1. Improvement in air quality

Graphs in figure 2 show the evolution of NO_2_, PM_2.5_, and CO in the cities of Casablanca and Marrakech before and during the quarantine. They also mention related Theil-Sen (TS) slopes and their significance (when in bold, they are significant). TS represents the magnitude of the trend during one day from the study period which counts 65 days. Multiplying this number of days by the TS gives the magnitude of the trend during the whole study period. TS assessment helps to validate the difference-in-difference estimate (DDE) presented in table 1. Overall, TS and DDE have the same order of magnitude.

**Table 1:** Air pollution changes due to the quarantine in Casablanca and Marrakech using DDE

|  | During vs before in 2020 |  | During vs before in 2016-2019 |  | Difference in difference |  |
| --- | --- | --- | --- | --- | --- | --- |
|  | Casablanca | Marrakech | Casablanca | Marrakech | Casablanca | Marrakech |
| **NO<sub>2</sub>** | -12.5 | -7.5 | -0.28 | -0.74 | -12.21 | -6.76 |
| **PM<sub>2.5</sub>** | -11.63 | -16.26 | 5.87 | -2.48 | -17.5 | -13.78 |
| **CO** | -0.06 | -0.21 | -0.02 | -0.1 | -0.04 | -0.12 |

**Figure 2:**
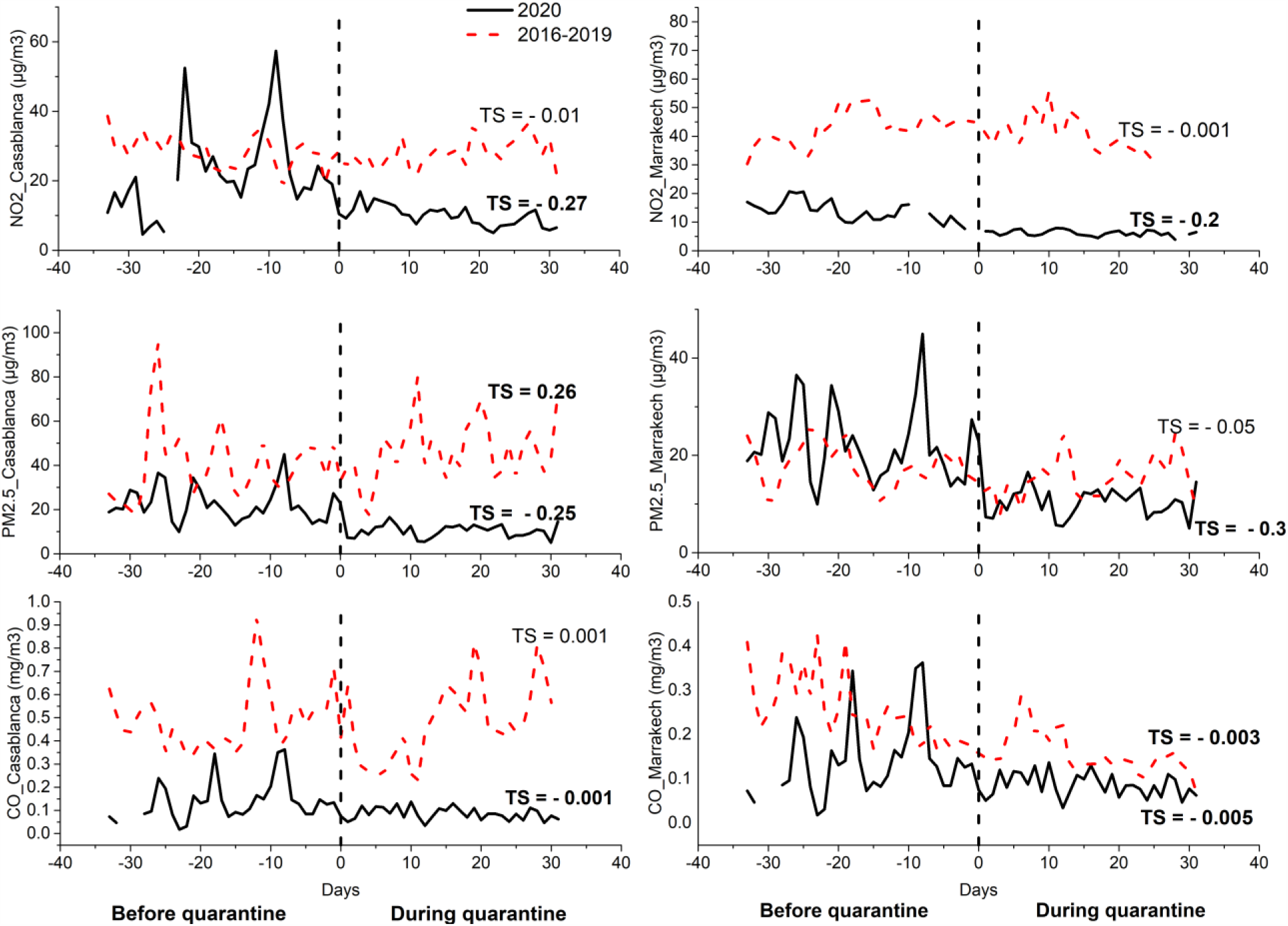
Air pollution changes due to the quarantine in Casablanca and Marrakech using TS **Bold character:** statitically significant

Hence, DDE shows that, because of the quarantine, NO_2_ dropped by -12 μg/m^3^ in Casablanca and -7 μg/m^3^ in Marrakech. PM_2·5_ dropped by -18 μg/m^3^ in Casablanca and -14 μg/m^3^ in Marrakech. CO dropped by -0.04 mg/m^3^ in Casablanca and -0.12 mg/m^3^ in Marrakech. All TS calculated slopes for the 2020 period are statistically significant and confirm the negative trends and order of magnitude of the DDE. TS slopes for the period 2016-2019 are not statistically significant.

These results are in complete agreement with the worldwide studies that confirmed the systematic link between COVID-19 lockdown and improvement in air quality levels.

### 3.2. COVID-19 vs air quality daily deaths

Figure 3 (a) and (b) show the cumulative number of deaths by COVID-19 in Morocco and the change in the number of daily mortalities, respectively. The results prove that the number of deaths increased between the 23^rd^ March and 4^th^ May and decreased notably since then. The rate of change in daily mortality decreases with time and reached the nil value by the 20^th^ of June.

**Figure 3:**
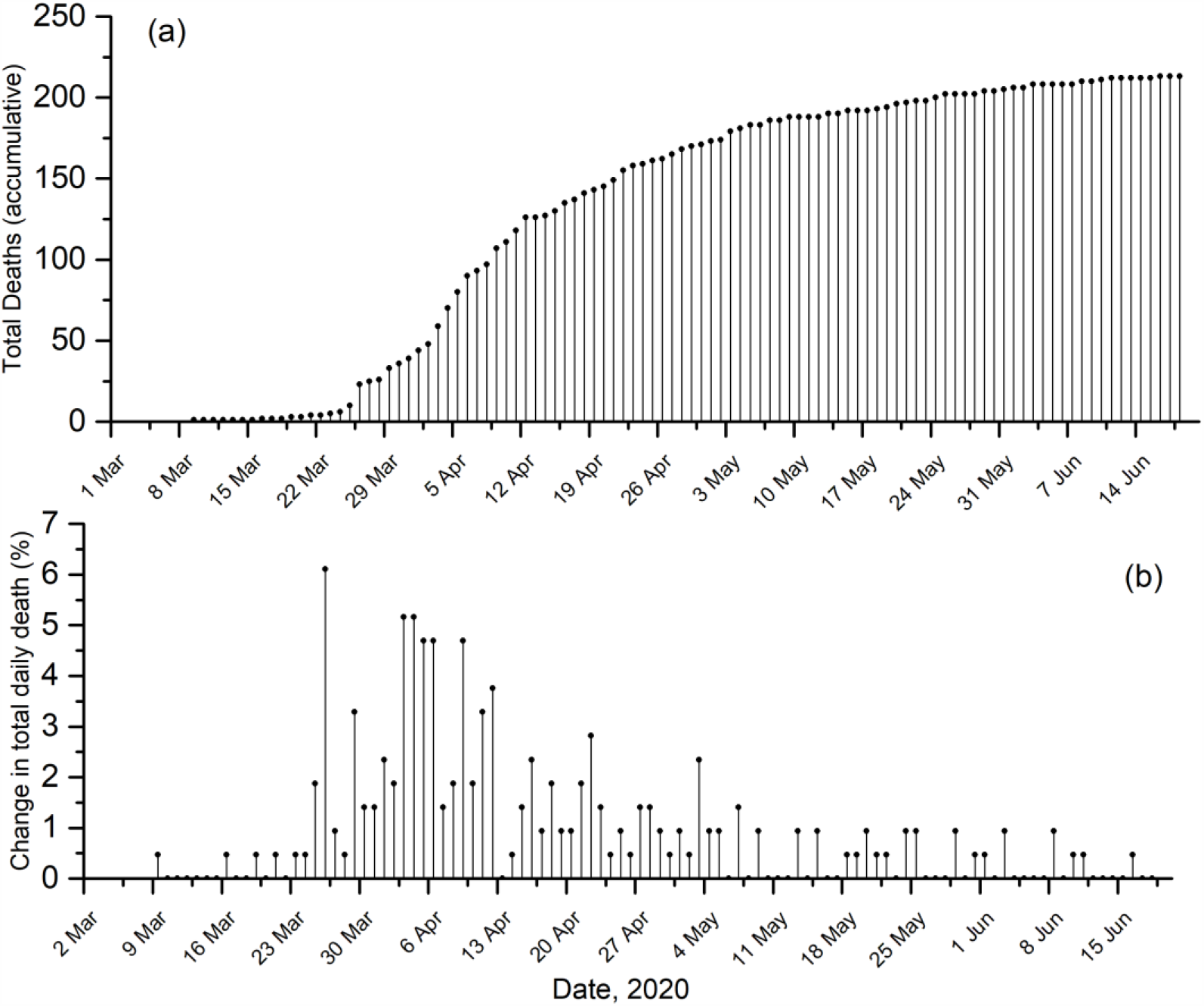
(a) Cumulative number of deaths by COVID-19 in Morocco (b) Change in the number of daily mortalities

According to the global report “Toxic Air: The Price of Fossil Fuels” of the international NGO Greenpeace MENA, Morocco is within a lengthy list of countries that suffer relatively from high estimated numbers of deaths annually due to air quality degradation. This number is estimated at 5,100 in 2018 (Farrow, A., Miller, K.A. & Myllyvirta, 2020). Dividing this value over 365 days helps obtain the daily average deaths due to air pollution and yields 14 deaths every day. This average fixed value was plotted against the daily reported deaths due to COVID-19 in Morocco (Fig. 4). Moreover, it is worth noting that outdoor air pollution by particulate matter death rate in Morocco is estimated at 63,1 per 100000 inhabitants (Ritchie, 2019). That of Covid-19 is of 0.59 per 100000 inhabitants as estimated by the Coronavirus resource center of the Johns Hopkins University (https://coronavirus.jhu.edu/data/mortality). All these results show a tremendous difference in deaths caused by COVID-19 and air pollution in Morocco.

**Figure 4:**
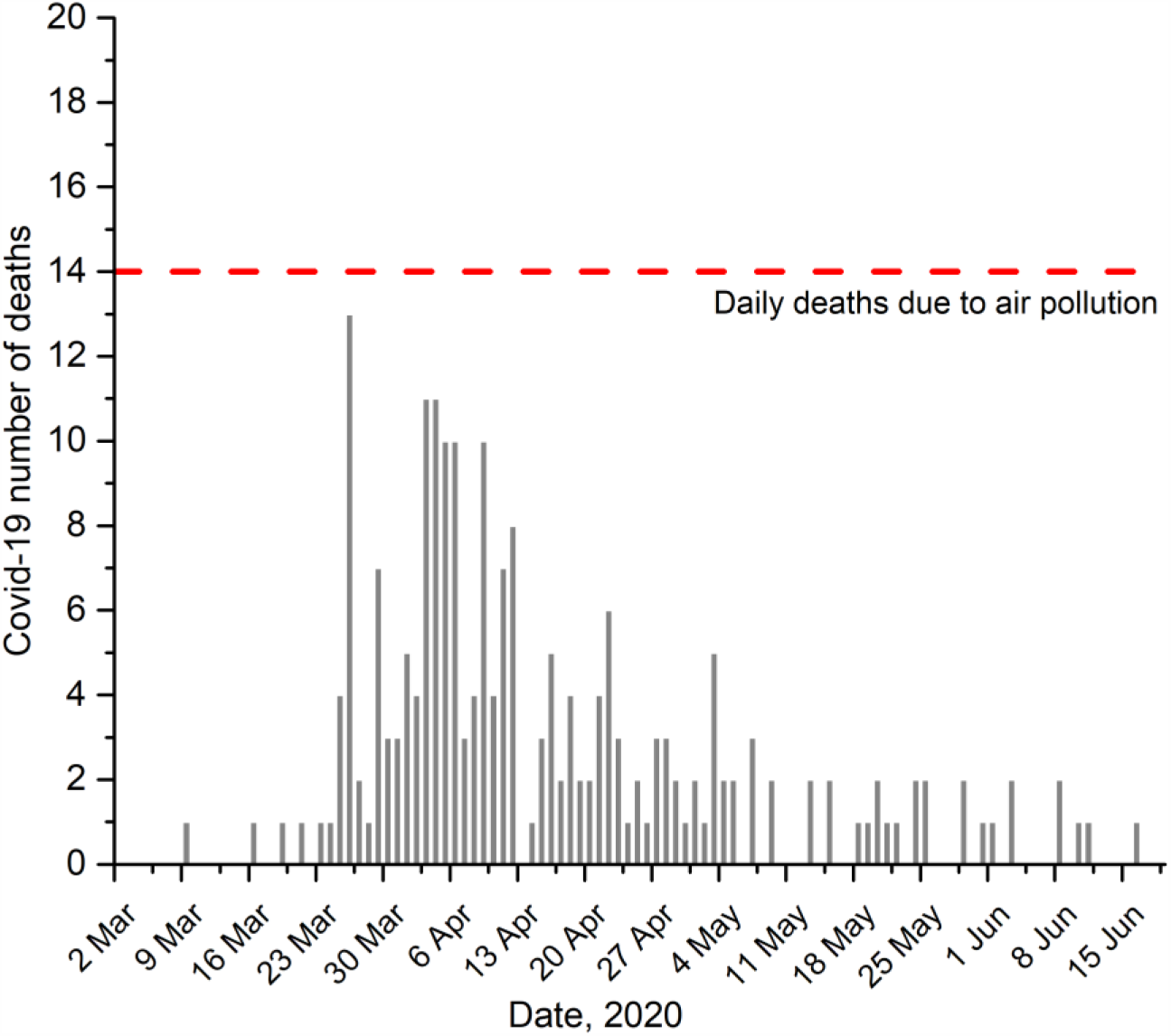
Daily deaths due to COVID-19 vs. averaged daily deaths due to poor air quality

### 3.3. Did COVID-19 save lives?

Table 2 shows the avoided cause-specific deaths due to air pollution reduction because of the COVID-19 home quarantine in Casablanca and Marrakech. We estimate that reduction in NO_2_ during the quarantine period avoided a total of 185 NO_2_ related deaths (95% CI 145–223) in Casablanca and 30 (95% CI 24–37) in Marrakech. Reduction in PM_2.5_ avoided a total of 48 PM_2.5_ related deaths (95% CI 70–89) in Casablanca and 15 (95% CI 10–19) in Marrakech. More than 60% of the avoidable deaths were from cardiovascular diseases. Cause-specific CRFs from single-pollutant models were used (Chen et al., 2020, 2017). Our results should be interpreted carefully because of the potential common impacts NO_2_ and PM_2·5_ may have on health. The risk of double counting is minor because, as suggested by the literature, effect estimates for NO_2_ and PM_2·5_ were similar between single-pollutant and two-pollutant models (Anenberg et al., 2010; Chen et al., 2018).

**Table 2:** Avoided cause-specific deaths (95% confidence interval) due to air pollution reduction because of the home quarantine in Casablanca and Marrakech

|  | Casablanca |  | Marrakech |  |
| --- | --- | --- | --- | --- |
|  | NO2 | PM2.5 | NO2 | PM2.5 |
| Total | 185(145,223) | 48(70,89) | 30(24,37) | 15(10,19) |
| Cardiovascular Disease | 96(76,126) | 45(30,59) | 16(12,21) | 10(6,13) |
| Hypertensive heart disease | 8(5,11) | 4(1,6) | 1(1,2) | 1(0,1) |
| Chronic respiratory diseases | 8(6,10) | 3(2,5) | 1(1,2) | 1(0,1) |
| Stroke | 23(13,30) | 4(2,5) | 9(5,13) | 2(1,3) |
| COPD | 5(7,9) | 2(3,4) | 1(1,1) | 1(0,1) |

Note: Home quarantine is the period between 20<sup>th</sup> March and 20<sup>th</sup> April 2020

Our findings imply that interventions to contain the COVID-19 outbreak led to improvements in air quality that brought health benefits in non-COVID-19 deaths, which could potentially have highly exceeded the current confirmed deaths attributable to COVID-19 in Morocco (229 deaths as of July 3, 2020).

## 4. Conclusion

In the present article, the effect of the imposed lockdown in Morocco in order to restrict the rapid spread of COVID-19 pandemic, on the air quality has been assessed based on NO_2_, PM_2.5_ and CO concentrations. Our estimates suggest that the undertaken lockdown led to improvements in air quality that brought health benefits in non-COVID-19 deaths. Specifically, our findings show the human health benefits related to cardiovascular diseases mortality that can be achieved if control measures are taken to reduce emissions from vehicles. Traffic restrictions or efforts to accelerate the transition to electric vehicles are good examples of these measures.

The results highlighted in the present work are not isolated and are in agreement with many other studies throughout the world. Thus, this paper and similar ones help to raise awareness about our responsibility towards the environment. It may also help to consider whether the COVID-19 lockdown scenario would be an efficient measure for preserving the environment and enhancing life quality in the urban ecosystem. However, cost effectiveness is one of the keys for policymakers to implement any control measure mainly that the lockdown has caused lower mobility and economic activity and shrinking the economy is not a sustainable solution to encounter the environmental challenge.

Moreover, the meteorological forcing is to take into consideration in concluding from such a study as they impact pollutants dispersion and concentration. Overall, the interlinkages between COVID-19, economy and climate are complex and depend on the climatic conditions, the duration of the emergency and reactions to it. Yet, like every crisis, the COVID-19 pandemic offers an important opportunity to draw lessons and to reconsider the way we treat the environment and the ecosystems.

## Data Availability

-COVID-19 statistics were collected from the official website set by the ministry of health of Morocco for that purpose (http://www.covidmaroc.ma/pages/Accueil.aspx)
- Air quality data were gathered from the National Weather Service in Morocco. Getting in touch with this organism is necessary to collect this type of data
- City-level demographics are obtained from the World Population Review webpage (https://worldpopulationreview.com/)
- Country-level air quality related mortality is collected from the Institute for Health Metrics and Evaluation (IHME)(https://vizhub.healthdata.org/gbd-compare/).

## Notes

### Competing Interest Statement

The authors have declared no competing interest.

### Funding Statement

The National Weather Service in Morocco provided air quality data

### Author Declarations

No IRB and/or ethics committee approvals were needed

